# SARS-CoV-2 infection in 86 healthcare workers in two Dutch hospitals in March 2020

**DOI:** 10.1101/2020.03.23.20041913

**Authors:** Marjolein F.Q. Kluytmans-van den Bergh, Anton G.M. Buiting, Suzan D. Pas, Robbert G. Bentvelsen, Wouter van den Bijllaardt, Anne J.G. van Oudheusden, Miranda M.L. van Rijen, Jaco J. Verweij, Marion P.G. Koopmans, Jan A.J.W. Kluytmans

**Affiliations:** Department of Infection Control, Amphia Hospital, Breda, the Netherlands; Amphia Academy Infectious Disease Foundation, Amphia Hospital, Breda, the Netherlands; Julius Center for Health Sciences and Primary Care, University Medical Center Utrecht, Utrecht, the Netherlands; Laboratory for Medical Microbiology and Immunology, Elisabeth-TweeSteden Hospital, Tilburg, the Netherlands; Department of Infection Control, Elisabeth-TweeSteden Hospital, the Netherlands; Microvida Laboratory for Microbiology, Bravis Hospital, Roosendaal, the Netherlands; Microvida Laboratory for Microbiology, Amphia Hospital, Breda, the Netherlands; Department of Medical Microbiology, Leiden University Medical Center, Leiden, the Netherlands; Department of Virology, Erasmus Medical Center, Rotterdam, the Netherlands

## Abstract

**Background:** On February 27, 2020, the first patient with COVID-19 was reported in the Netherlands. During the following weeks, nine healthcare workers (HCWs) were diagnosed with COVID-19 in two Dutch teaching hospitals, eight of whom had no history of travel to China or Northern-Italy. A low-threshold screening regimen was implemented to determine the prevalence and clinical presentation of COVID-19 among HCWs in these two hospitals.

**Methods:** HCWs who suffered from fever or respiratory symptoms were voluntarily tested for SARS-CoV-2 by real-time reverse-transcriptase PCR on oropharyngeal samples. Structured interviews were conducted to document symptoms for all HCWs with confirmed COVID-19.

**Findings:** Thirteen-hundred fifty-three (14%) of 9,705 HCWs employed were tested, 86 (6%) of whom were infected with SARS-CoV-2. Most HCWs suffered from relatively mild disease and only 46 (53%) reported fever. Eighty (93%) HCWs met a case definition of fever and/or coughing and/or shortness of breath. None of the HCWs identified through the screening reported a travel history to China or Northern Italy, and 3 (3%) reported to have been exposed to an inpatient known with COVID-19 prior to the onset of symptoms.

**Interpretation:** Within two weeks after the first Dutch case was detected, a substantial proportion of HCWs with fever or respiratory symptoms were infected with SARS-CoV-2, probably caused by acquisition of the virus in the community during the early phase of local spread. The high prevalence of mild clinical presentations, frequently not including fever, asks for less stringent use of the currently recommended case-definition for suspected COVID-19.

**RESEARCH IN PERSPECTIVE:** *Evidence before this study:* This study was conducted in response to the global spread of SARS-CoV-2, and the detection of eight healthcare workers (HCWs) in two Dutch teaching hospitals within two weeks after the first patient with COVID-19 was detected in the Netherlands who had no history of travel to China or Northern-Italy, raising the question of whether undetected community circulation was occurring.

*Added value of this study:* To the best of our knowledge, this report is the first to describe the prevalence, the clinical presentation and early outcomes of COVID-19 in HCWs, which may be helpful for others seeking to identify HCWs suspected for COVID-19 in an outbreak situation.

*Implications of all the available evidence:* We describe that within two weeks after the first Dutch case was detected, a substantial proportion of HCWs with fever or (mild) respiratory symptoms were infected with SARS-CoV-2, probably caused by acquisition of the virus in the community during the early phase of local spread. The high prevalence of mild clinical presentations, frequently not including fever, asks for less stringent use of the currently recommended case-definition for suspected COVID-19.

## INTRODUCTION

Since December 2019, the world has been in the grip of the severe acute respiratory syndrome coronavirus 2 (SARS-CoV-2) and the disease it causes, coronavirus disease 2019 (COVID-19).^1^ On February 27, 2020, the first patient with COVID-19 was detected in the Netherlands, linked to a trip to Northern-Italy between February 18, 2020 and February 21, 2020.^2^ During the following weeks, more cases of COVID-19 were identified in the Netherlands, including nine healthcare workers (HCWs) in two Dutch teaching hospitals in the southern part of the Netherlands who were diagnosed between March 2, 2020 and March 6, 2020. Eight of these nine healthcare workers (HCWs) had no history of travel to China or Northern-Italy, raising the question of whether undetected community circulation was occurring. As these findings coincided with the seasonal influenza peak,^3^ and SARS-CoV-2 infection in HCWs could lead both to sick leave and introduction of the virus into the hospitals, this finding prompted a demand for testing HCWs. Following initial observations of SARS-CoV-2 detection in persons with mild symptoms not meeting the definition for case finding,^1^ a low-threshold screening regimen was implemented to determine the prevalence and the clinical presentation of COVID-19 among HCWs in these two hospitals.

## METHODS

### Study design

A cross-sectional study with short-term follow-up was conducted in two teaching hospitals (700-bed Amphia Hospital, Breda, the Netherlands; 800-bed Elisabeth-TweeSteden Hospital, Tilburg, the Netherlands), employing 9,705 HCWs, 18% of whom are male (Figure 1).

**Figure 1.**
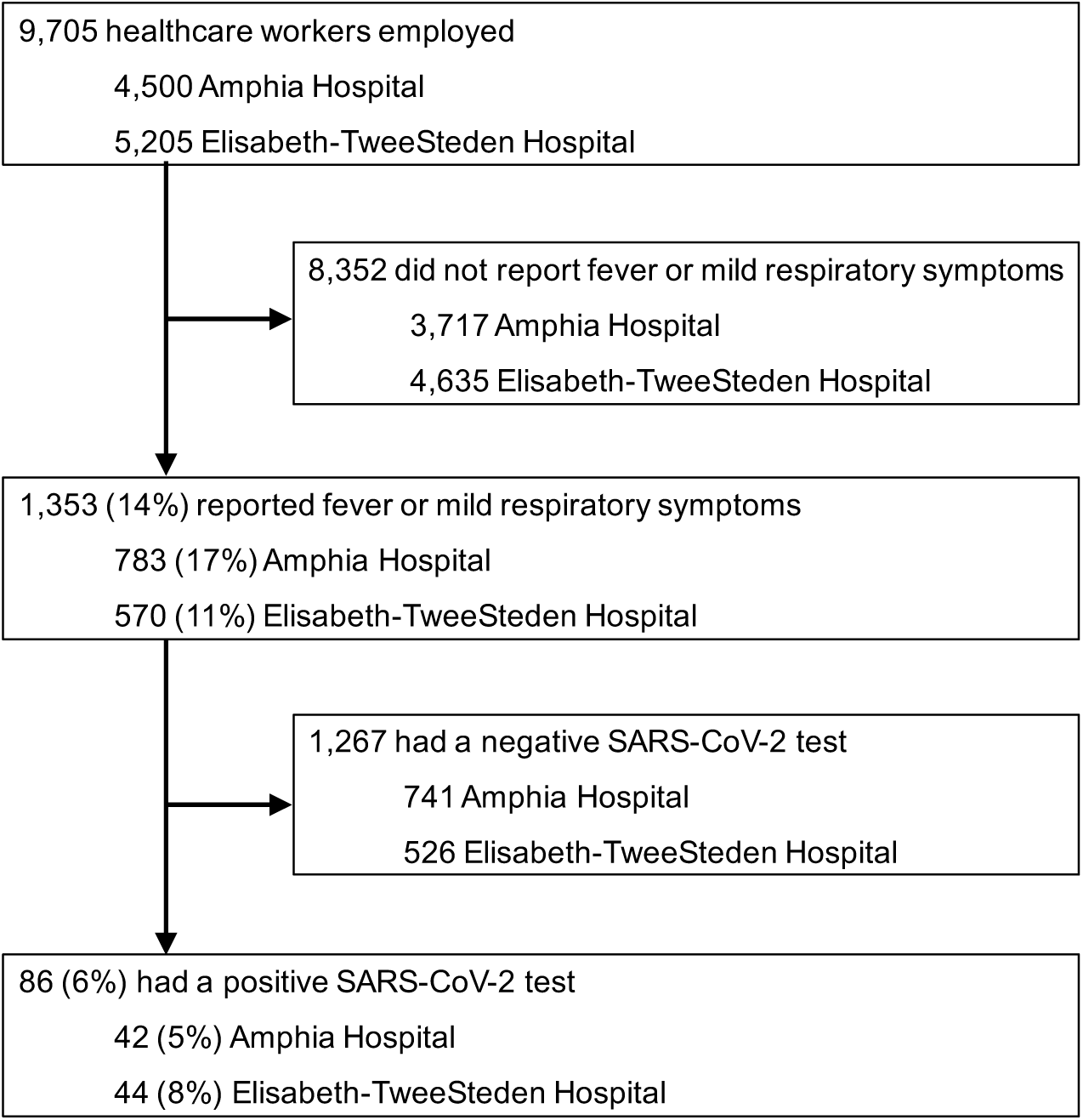
Recruitment of healthcare workers in two Dutch hospitals in the southern part of the Netherlands, March 2020.

The study was reviewed by the Ethics Committee Brabant, the Netherlands (METC Brabant/20.134/NW2020-26). The study was judged to be beyond the scope of the Medical Research Involving Human Subjects Act and a waiver of written informed consent was granted. Verbal informed consent was obtained from all HCWs for SARS-CoV-2 testing, from SARS-CoV-2 infected HCWs for data collection. Data were de-identified before analysis.

### Study population

Between March 7, 2020 and March 12, 2020, HCWs in both teaching hospitals who suffered from fever or (mild) respiratory symptoms in the last ten days were tested voluntarily for SARS-CoV-2 infection, in accordance with the local infection control policy during outbreaks.

### Procedures

A semi-quantitative real-time reverse-transcriptase PCR (RT-PCR, 45 cycles) targeting the E-gene was performed on self-collected oropharyngeal samples as described previously (Appendix).^4^ Structured interviews were conducted between March 12, 2020 and March 16, 2020 to document symptoms for all HCWs with confirmed COVID-19, including those diagnosed before March 7, 2020 (Appendix). Recovery was defined as being without symptoms for more than 24 h.

### Statistical analysis

Given the descriptive nature of the report sample size calculations and analysis for statistical significance were not performed. Continuous variables were expressed as medians and ranges. Categorical variables were summarised as counts and percentages. There were no missing data. All analyses were performed with SPSS version 25.0 (IBM, Armonk, NY, USA).

## RESULTS

A total of 1,353 (14%) HCWs were screened, 86 (6%) of whom were infected with SARS-CoV-2 (Figure 1). HCWs with COVID-19 were employed in 52 different hospital departments, including 36 medical wards, had a median age of 49 years (range 22-66 years) and 15 (17%) were male (Table 1). Most HCWs with COVID-19 suffered from relatively mild disease. Forty-six (53%) HCWs had fever during the course of illness, another 10 (12%) reported a feverish feeling without having measured their temperature. Eighty (93%) HCWs met a case definition of fever and/or coughing and/or shortness of breath. Extending this case definition with severe myalgia and/or general malaise would capture all 86 (100%) HCWs with COVID-19 in this evaluation. Other frequent symptoms were headache, a runny nose, a sore throat, chest pain, diarrhea and loss of appetite. Seven (8%) indicated that they were already symptomatic before February 27, 2020, the day the first Dutch patient with COVID-19 was diagnosed (Figure 2). Four (5%) HCWs had recovered on the day of screening, 19 (22%) on the day of the interview, with a median duration of illness of 8 days (range 1-20 days) (Table 1). Two (2%) HCWs were admitted to the hospital and did not develop critical disease up to the moment of reporting. Coughing, shortness of breath, general malaise, loss of appetite and altered or loss of taste were more frequent in HCWs who were interviewed during the second week of illness. Three (3%) HCWs reported to have been exposed to an inpatient known with COVID-19 prior to the onset of symptoms, and 54 (63%) mentioned to have worked while being symptomatic.

**Table 1.**
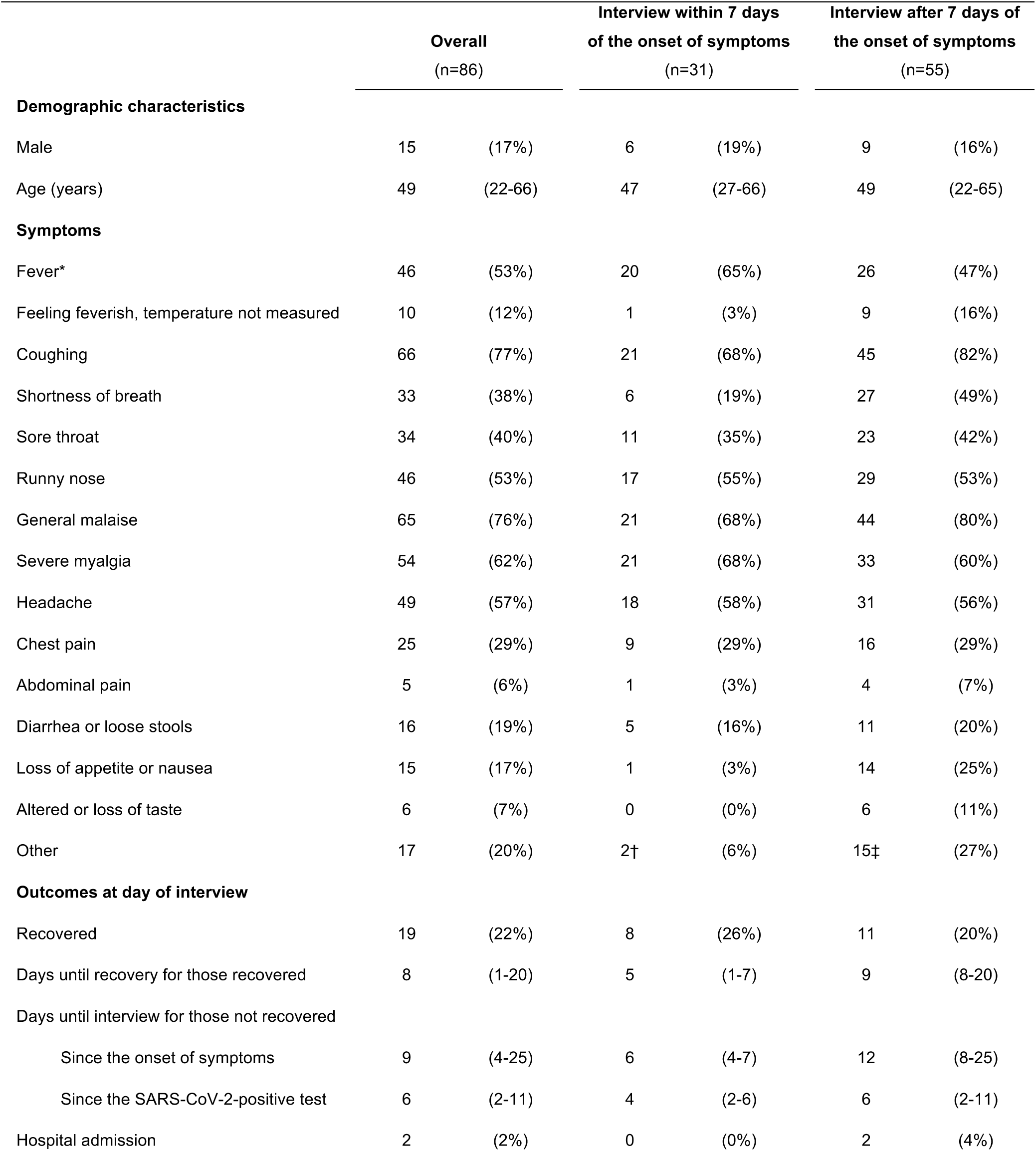
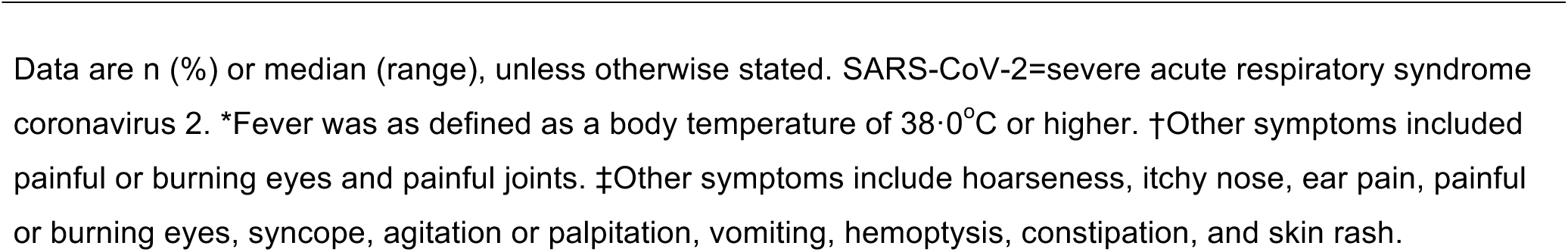
Demographic characteristics, symptoms during the course of illness and outcomes of healthcare workers with confirmed coronavirus disease 2019.

|  | Overall<br>(n=86) |  | Interview within 7 days<br>of the onset of symptoms<br>(n=31) |  | Interview after 7 days of<br>the onset of symptoms<br>(n=55) |  |
| --- | --- | --- | --- | --- | --- | --- |
| Demographic characteristics |  |  |  |  |  |  |
| Male | 15 | (17%) | 6 | (19%) | 9 | (16%) |
| Age (years) | 49 | (22-66) | 47 | (27-66) | 49 | (22-65) |
| Symptoms |  |  |  |  |  |  |
| Fever* | 46 | (53%) | 20 | (65%) | 26 | (47%) |
| Feeling feverish, temperature not measured | 10 | (12%) | 1 | (3%) | 9 | (16%) |
| Coughing | 66 | (77%) | 21 | (68%) | 45 | (82%) |
| Shortness of breath | 33 | (38%) | 6 | (19%) | 27 | (49%) |
| Sore throat | 34 | (40%) | 11 | (35%) | 23 | (42%) |
| Runny nose | 46 | (53%) | 17 | (55%) | 29 | (53%) |
| General malaise | 65 | (76%) | 21 | (68%) | 44 | (80%) |
| Severe myalgia | 54 | (62%) | 21 | (68%) | 33 | (60%) |
| Headache | 49 | (57%) | 18 | (58%) | 31 | (56%) |
| Chest pain | 25 | (29%) | 9 | (29%) | 16 | (29%) |
| Abdominal pain | 5 | (6%) | 1 | (3%) | 4 | (7%) |
| Diarrhea or loose stools | 16 | (19%) | 5 | (16%) | 11 | (20%) |
| Loss of appetite or nausea | 15 | (17%) | 1 | (3%) | 14 | (25%) |
| Altered or loss of taste | 6 | (7%) | 0 | (0%) | 6 | (11%) |
| Other | 17 | (20%) | 2† | (6%) | 15‡ | (27%) |
| Outcomes at day of interview |  |  |  |  |  |  |
| Recovered | 19 | (22%) | 8 | (26%) | 11 | (20%) |
| Days until recovery for those recovered | 8 | (1-20) | 5 | (1-7) | 9 | (8-20) |
| Days until interview for those not recovered |  |  |  |  |  |  |
| Since the onset of symptoms | 9 | (4-25) | 6 | (4-7) | 12 | (8-25) |
| Since the SARS-CoV-2-positive test | 6 | (2-11) | 4 | (2-6) | 6 | (2-11) |
| Hospital admission | 2 | (2%) | 0 | (0%) | 2 | (4%) |

Data are n (%) or median (range), unless otherwise stated. SARS-CoV-2=severe acute respiratory syndrome coronavirus 2. \*Fever was as defined as a body temperature of 38·0°C or higher. †Other symptoms included painful or burning eyes and painful joints. ‡Other symptoms include hoarseness, itchy nose, ear pain, painful or burning eyes, syncope, agitation or palpitation, vomiting, hemoptysis, constipation, and skin rash.

**Figure 2.**
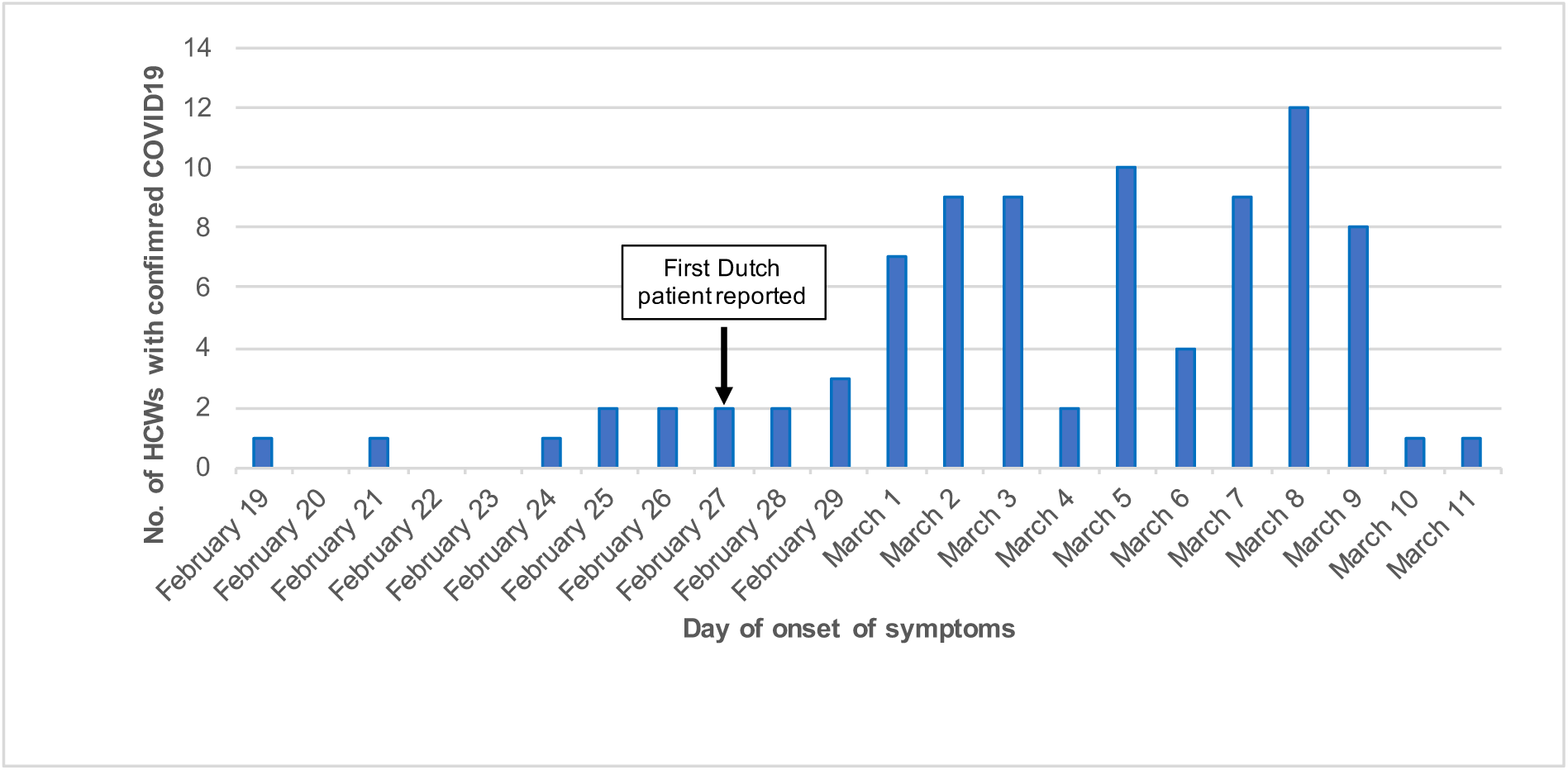
Day of onset of symptoms for 86 healthcare workers with confirmed coronavirus disease 2019 in two hospitals in the southern part of the Netherlands, March 2020. HCW=healthcare worker.

The median RT-PCR Ct value was 27·0 (range 14·5-38·5). Within the limited resolution in time since the onset of symptoms, Ct values tended to be higher in HCWs who were tested later in the course of disease (Figure 3). Ct values were similar for HCWs with and without fever on the day of screening (median 25·1 and 27·6, respectively), and for HCWs with and without any symptoms on the day of screening (median 27·0 and 26·7, respectively).

**Figure 3.**
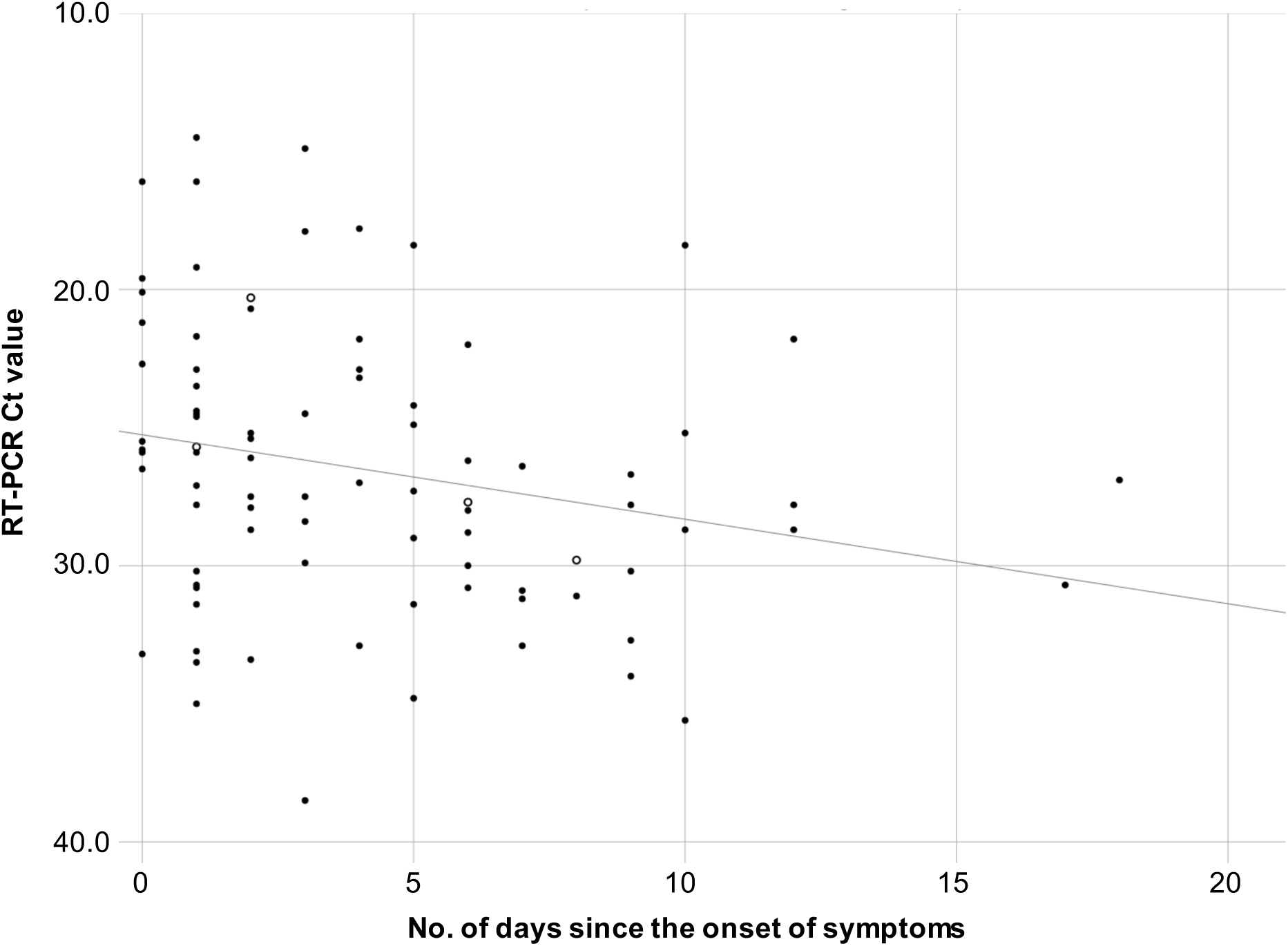
Cycle threshold values for the semi-quantitative reverse-transcriptase PCR (E-gene) on self-collected oropharyngeal samples from health care workers in two Dutch hospitals in the southern part of the Netherlands (March 2020) in relation to the number of days since the onset of symptoms. Open circles indicate Ct values for HCWs who had recovered on the day of screening. Ct=cycle threshold; PCR=polymerase chain reaction. RT=reverse-transcriptase.

## DISCUSSION

Two weeks after the first Dutch patient with COVID-19 was reported, the prevalence of COVID-19 in HCWs with fever or respiratory symptoms in two Dutch hospitals in the southern part of the Netherlands was 6%. This unexpected high prevalence supported the hypothesis of hidden community spread of SARS-CoV-2 and is considered a minimal estimate of the prevalence in all HCWs at the time of screening. Only HCWs with (recent) symptoms were screened, and oropharyngeal swabs were used for testing, which may have a slightly lower sensitivity than a nasopharyngeal swab.^5^ Another possible explanation for the unexpectedly high prevalence would be hospital-acquisition. However, all patients with fever or respiratory symptoms in both hospitals were routinely tested for SARS-CoV-2. At that time, a limited number of infected patients was nursed under strict isolation precautions, and only three SARS-CoV-2-infected HCWs mentioned exposure to an inpatient known with COVID-19. There was no clustering of infected HCWs in specific departments. The low percentage of males among HCWs with COVID-19 (17%) reflects that among the source population of HCWs in the two participating hospitals (18%).

Most HCWs suffered from mild disease as compared to the clinical presentation and outcomes reported for hospitalised patients so far.^6,7^ Notably, fever or a feverish feeling was frequently not reported. A question is what is a sensitive case definition for early detection of SARS-CoV-2 infected individuals. At the time of the study, the internationally recommended case definition included a history of travel to China or Northern-Italy,^1^ which did not apply for any of the infected HCWs identified through our screening. When using the definition without travel history to capture community transmission, about 40% of HCWs with COVID-19 in our hospitals still would not have been detected. Sensitive detection of COVID-19 cases in HCWs is crucial for hospital infection prevention policy, particularly for those who work with vulnerable patients. We therefore suggest adjusting the currently used case-definition for suspected COVID-19 in HCWs by taking fever as one of the possible symptoms and not as a required symptom. Further improvement of the sensitivity of COVID-19 detection in HCWs can be achieved by adding severe myalgia and general malaise to the case-definition.

To the best of our knowledge, this report is the first to describe the prevalence, the clinical presentation and early outcomes of COVID-19 in HCWs, which may be helpful for others seeking to identify HCWs suspected for COVID-19 in an outbreak situation. A limitation of our evaluation is that screening of HCWs was based on the presence of fever or mild respiratory symptoms in the last ten days, and that no data were collected in HCWs without these symptoms. The sensitivity and specificity of the reported symptoms could therefore not be estimated.

In conclusion, during the containment phase and within two weeks after the first Dutch case was detected, a substantial proportion of HCWs with fever or respiratory symptoms were infected with SARS-CoV-2, probably caused by acquisition of the virus in the community during the early phase of local spread. This observation confirms the insidious nature of SARS-CoV-2 spread, given the high prevalence of mild clinical presentations that may go undetected.^8^ The spectrum of relatively mild symptoms present in HCWs with COVID-19, frequently not including fever, asks for less stringent use of the currently recommended case-definition for suspected COVID-19.

## Data Availability

All data analysed are available from the presenting author on reasonable request.

## Contributors

SP, JV and MKo were involved in laboratory testing. MKl, AB, RB, WB, AO, MR and JK were involved in data collection. MK analyzed and interpreted the data and drafted the manuscript. All authors revised the manuscript and read and approved the final version.

## Data sharing

All data collected and analyzed for this report are available from the first author on reasonable request.

## Acknowledgments

We are grateful to the healthcare workers of the Amphia Hospital (Breda, the Netherlands) and the Elisabeth-TweeSteden Hospital (Tilburg, the Netherlands) for participating in the low-threshold screening program. We thank infection control practitioners and microbiology technicians of the participating hospitals and laboratories for their contribution in the collection of epidemiological and microbiological data. MKo participates in the RECOVER project that is funded by the European Commission under H2020 call SC1-PHE-CORONAVIRUS-2020. The funder had no role in study design, data collection, data analysis, data interpretation, or writing of the report. The corresponding author had full access to all data in the study and had final responsibility for the decision to submit for publication.

## APPENDIX

### Semi-quantitative real-time reverse-transcriptase PCR for SARS-CoV-2

After an external lysis step (1:1 with lysis/binding buffer (Roche Diagnostics, the Netherlands), total nucleic acids were extracted using MagnaPure96 (Roche) with an input volume of 500 µl and output volume of 100 µl. The extraction was internally controlled by the addition of a known concentration of phocine distemper virus (PDV).^1^ Subsequently 10 μl extracted nucleic acids was amplified in three singleplex reactions 25 μl final volume, using TaqMan Fast Virus 1-Step Master Mix (Thermofisher, Nieuwerkerk a/d IJssel, the Netherlands), and 1 μl of primers and probe mixture for E gene, RdRp gen as described previously.^2^ Amplification was performed in a 7500SDS (Thermofisher) with a cycling profile of 5 min at 50°C, 20 s at 95°C, 45 cycles of 3 s at 95°C and 30 s at 58°C. Alternatively, total nucleic acids were extracted, with a known concentration of PDV as internal control, using the QIAsymphony DSP virus/pathogen midi kit and pathogen complex 400 protocol of the QIAsymphony Sample Processing (SP) system (Qiagen, Hilden, Germany) with an input volume 400 µl and output volume of 110 µl. Amplification reactions were performed in a volume of 25 µL with TaqMan^®^ Fast Virus 1-Step Master Mix (Thermofisher) and 10 µL extracted nucleic acids. A duplex PCR for E-gen/PDV and if positive a duplex PCR for RdRP/PDV with optimised primer and probe concentrations were performed.^1,3^ Amplification using Rotorgene (QIAgen) consisted of 5 min at 50°C, 15 min at 95°C followed by 45 cycles of 15 s at 95°C, 30 s at 60°C, and 15s at 72°C. Validations of RT-PCR procedures were performed according to International Standards Organization guidelines 15189 (http://www.iso.org/iso/search.htm).

## Questionnaire used for healthcare workers with COVID-19

1. Hospital
2. Department
3. Position
4. Sex
5. Year of birth
6. Symptoms on day of SARS-CoV-2 testing
  a. Fever (>=38 °C)
  b. Coughing
  c. Sore throat
  d. Runny nose
  e. (Severe) myalgia
  f. General malaise
  g. Headache
  h. Shortness of breath
  i. Chest pain
  j. Abdominal pain
  k. Diarrhea
  l. Other, please specify
7. Symptoms in 10 days before the day of SARS-CoV-2 testing
  a. Fever (>=38 °C)
  b. Coughing
  c. Sore throat
  d. Runny nose
  e. (Severe) myalgia
  f. General malaise
  g. Headache
  h. Shortness of breath
  i. Chest pain
  j. Abdominal pain
  k. Diarrhea
  l. Other, please specify
8. Symptoms between day of SARS-CoV-2 testing until day of interview
  a. Fever (>=38 °C)
  b. Coughing
  c. Sore throat
  d. Runny nose
  e. (Severe) myalgia
  f. General malaise
  g. Headache
  h. Shortness of breath
  i. Chest pain
  j. Abdominal pain
  k. Diarrhea
  l. Other, please specify
9. Hospital admission
10. Having worked while being symptomatic
11. First day of symptoms
12. Last day of symptoms (if recovered)
13. Day of SARS-CoV-2 test
14. Day of interview

